# Application of pooled testing in screening and estimating the prevalence of COVID-19

**DOI:** 10.1101/2020.05.26.20113696

**Authors:** Pritha Guha, Apratim Guha, Tathagata Bandyopadhyay

## Abstract

The recent emergence of the COVID-19 pandemic has posed an unprecedented healthcare challenge and catastrophic economic and social consequences to the countries across the world. The situation is even worse for emerging economies like India. WHO recommends mass scale testing as one of the most effective ways to contain its spread and fight the pandemic. But, due to the high cost and shortage of test kits, specifically in India, the testing is restricted to only those who are symptomatic. In this context, pooled testing is recommended by some experts as a partial solution to overcome this problem. In this article, we explain the basic statistical theory behind the pooled testing procedure for screening as well as prevalence estimation. In real world situations, the tests are imperfect, and lead to false positive and false negative results. We provide theoretical explanation of the impact of these diagnostic errors on the performances of individual testing and pooled testing procedures. Finally, we study the effect of misspecification of sensitivity and specificity of tests on the estimate of prevalence, an issue, which is debated a lot among the scientists in the context of COVID-19. Our theoretical investigations lead to some interesting and precise understanding of some of these issues.

## 1 Introduction

Since its detection in the Wuhan province of China at the end of December 2019, the COVID-19 pandemic, caused by the virus SARS-CoV-2, has created catastrophic social, economic and health consequences for countries around the world. For an emerging economy like India, it poses a formidable challenge to the policy makers to contain its spread at a level so that the health care infrastructure is not overwhelmed. At the time of writing this article, in India, the number infected is around 150,000 and more than 4300 deaths have occurred due to COVID-19. A large part of the country is under ‘lockdown’. Given its unprecedented economic and social consequences, the policy makers are now desperately looking for an exit route to bring the economy back to normalcy. Effective implementation of social distancing, mass testing, contact tracing and quarantining is essential to contain the spread of the virus. Given that a large percentage of COVID-19 cases are asymptomatic, and are responsible for spreading the virus^1-3^, it has been advised by the World Health Organization (WHO) that the most effective way to control the spread of the disease is to test as many people as possible. This was found to be effective in containing the spread of the virus in countries like South Korea, Singapore and China^4-6^. However, the high cost and the short supply of testing kits^7^ are the main impediments for a country like India in conducting mass testing. In order to ramp up the testing efforts, some experts^8-11^ are recommending the use of pooled testing technique, a technique originally proposed by Dorfman^12^ to increase the speed, and reduce the cost, of screening the US army recruits for syphilis during the Second World War. Recently the Indian Council for Medical Research (ICMR) has given its approval^13^ and issued detailed guidelines^14^ for carrying out pooled testing for screening purposes.

It is interesting to observe that most experts recommend use of pooled testing only for the screening purpose. However, the data collected by pooled testing can also be effectively utilized for the estimation of prevalence of the virus, which is a later development in pooled testing research^15^. This fact is not as widely known as the use of pooled testing for screening purpose in the general scientific community. As observed by many^16,17^, the estimation of prevalence of the SARS-CoV-2 virus is crucial for an accurate estimation of fatality rate. The initial estimate of fatality rate given by WHO ^18^ was scandalously high, because it was based only on deaths among symptomatic patients. Later, it was revised downward after taking into consideration the fact that a significant percentage of the infected population is asymptomatic, and hence remain undetected. If 80% of the cases are asymptomatic, then the fatality rate computed only from the symptomatic cases would be five times higher than the true fatality rate. Such misinformation may create an unnecessary panic in the minds of the policy makers, which may lead to wrong decisions. Recently, a few planned serology surveys were carried out for the estimation of the prevalence of the SARS-CoV-2 virus (e.g., Sempos and Tian^19^), and the results suggest that the overestimation of fatality rate in the population may be between fifty to eighty five times of the original fatality rate. These results created waves in the scientific community. We will revisit these results in Section 4.

This article mainly serves two purposes. First, we present the statistical theory of the pooled testing procedure for screening, and also for the estimation of prevalence using basic probability theory^20^. Next, we discuss some practical issues arising out of the fact that these tests are imperfect. In other words, the tests may yield false positive and/or false negative results. Naturally, it raises an important practical question: what is the impact of these diagnostic errors on screening as well as on prevalence estimation? We study these effects using some hypothetical scenarios and discuss its implications in the context of COVID-19 pandemic.

The rest of the article is divided as follows. In Section 2 we present the statistical theory of pooled testing. In Section 3 we discuss the consequences when the test is imperfect. In Section 4, we discuss the estimation of prevalence using pooled data, and compare it with the estimation procedure with individual data. Finally, we conclude with some remarks in Section 5.

## 2 Dorfman pooled testing technique

Pooled testing was originally introduced by Dorfman^12^ in infectious disease studies to reduce the cost and increase the speed of data collection. Typically, for an infectious disease, a sample of blood or urine is tested for presence or absence of the disease. Rarer the disease, more effective is the pooled testing technique. Dorfman’s technique runs as follows. Suppose *n* samples of blood are to be tested for presence or absence of a disease. A sample is either positive (disease present) or negative (disease absent). We attach a Bernoulli variable *Y* to each sample. We assume *Y* =1 if the sample is positive, and *Y* = 0 if it is negative. Instead of testing the samples individually, the samples are pooled into *J* groups of sizes *n*_1_, …,*n_J_*. Let *Y_ij_* denote the *Y*-value for the *i*-th sample in the *j*-th group. The samples in each group are pooled and tested. Instead of observing *Y_ij_*, in the pooled testing set-up, we observe 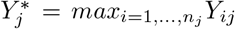. It is further assumed that, 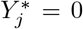, if and only if all samples in the *j*-th group are negative, i.e., *Y_ij_* = 0, *i* = 1, …, *n_j_* and 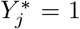, if and only if at least one sample in the *j*-th group is positive, i.e., *Y_ij_* = 1, for at least one *i* = 1, *..,n_j_*. If for a group, the test outcome is positive, then the samples in the group are individually tested to detect the positive samples, and hence the diseased individuals. If the result is negative for a group then no further testing is required. Thus, for a positive test outcome, the number of tests required is one more than the group size, and for a negative test outcome, a single test is enough. If the chance of a positive test outcome for a group is small, in other words, the prevalence of the disease is low, then the average number of tests required would be much smaller in a group testing set-up than individual testing. Thus, it leads to considerable savings in the cost and time of testing. The process diagram is presented in Figure 1 for a group of size *k*.

**Figure 1:**
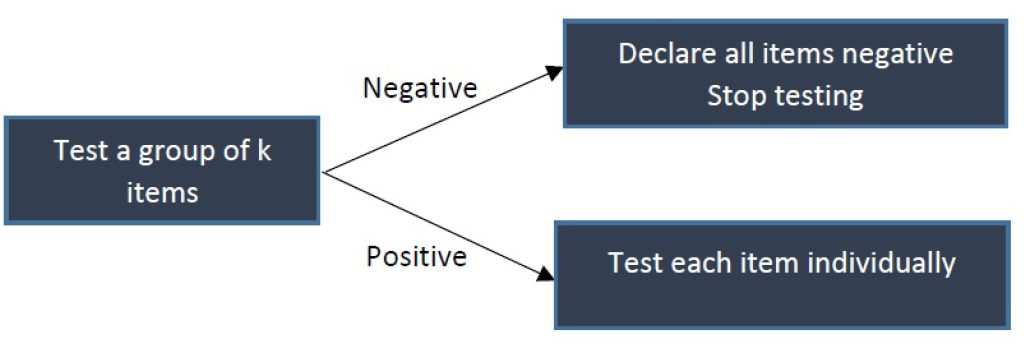
Process diagram of Dorfman’s algorithm for pooled testing

Now, suppose that the prevalence of a disease is *p*, and assume without loss of any generality *n_j_* = *k*, i.e., all groups are of equal size, and consequently *n* = *Jk*. Notice that, 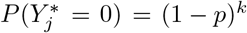 and 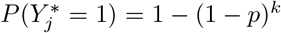. Further, suppose *N_j_* denotes the number of tests for the *j*-th group. Clearly, it is equal to *k* + 1 if 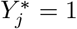, and 1 if 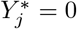. Thus the expected number of tests for the *j*-th group is

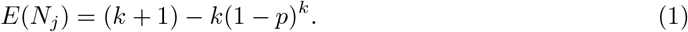

If *p* is small then *E*(*N_j_*) is close to 1 even for a sufficiently large group size *k*, and thus leading to a substantial saving in time and cost than individual testing. Notice that the technique is mainly useful when the prevalence is low.

The simplicity of the idea, and its easy implementation, has led to its wide applicability in different fields (see Roy and Banerjee^15^ and the references therein). Its use in the screening of sexually transmitted disease like chlamydia, gonorrhoea^21^ and HIV^22,23^ is worth mentioning. American Red Cross and the European Blood Alliance routinely use it to screen donated blood for infectious diseases^24,25^. Given the huge shortage of test kits around the world, researchers worldwide are recommending the use of pooled testing for COVID-19^8,26-28^.

## 3 Pooled testing for screening when the test is imperfect

Dorfman^12^ proposed the pooled testing technique assuming the test to be perfect, i.e., the chance of a false positive or a false negative test result to be zero. However, in reality, tests are far from perfect. In testing for the SARS-CoV-2 virus, two types of tests are mainly in use. “The first is a reverse-transcription polymerase chain reaction test, or RT-PCR. This is the most common diagnostic test used to identify people currently infected with SARS-CoV-2. It works by detecting viral RNA in a person’s cells – most often collected from their nose. The second test being used is called a serological or antibody test. This test looks at a person’s blood to see if they have produced antibodies for the SARS-CoV-2 virus. If a test finds these antibodies, it means a person was infected and made antibodies in response” ^29^. How accurate are these tests? The accuracy of a test is measured by two numbers: sensitivity and specificity attached to it. If a test has 5% chance of a false negative (positive) outcome, its sensitivity (specificity) is 95%. Although in laboratory setting it is observed that RT-PCR test has high sensitivity and specificity, in the real world testing condition the sensitivity and the specificity are usually much lower because the testing conditions, the method of sample collection, and the sample preservation technique are far from perfect. For example, in real world testing condition the sensitivity of RT-PCR test ranges between 66% to 80%^29^. Thus, out of three infected people on the average one may test negative. The sensitivity and specificity of the antibody tests are also found to be very high in laboratory setting, but, then again, in real world condition the accuracy is bound to suffer.

Using the same notations as above, we now find *E*(*N_j_*) assuming that the sensitivity and specificity of the test are *S_e_* and *S_p_*, respectively. Notice that for the *j*-th group, we now observe 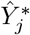 instead of 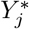, a surrogate (or proxy) for 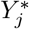, where 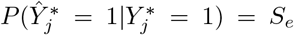 and 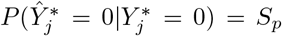. A simple probability calculation then yields 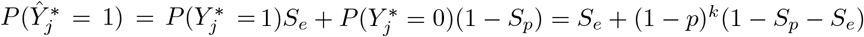, and hence,

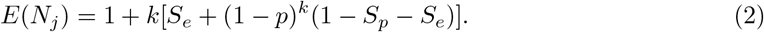

The expected number of tests for *n* people would then be *J* × *E*(*N_j_*), and hence compared to the individual testing, the reduction in the number of tests achieved by using group testing is *J*(*k* - *E*(*N_j_*)). This reduction is a measures of the *efficiency* of the pooled testing procedure. Also, notice that for *S_p_* = *S_e_* = 1, Equation (2) reduces to (1).

For a perfect test, there is no chance of misclassification of an infected as non-infected or vice versa, and hence the efficiency of pooled testing procedure is measured only by *E*(*N_j_*). However, for an imperfect test, *E*(*N_j_*) is not enough to judge the efficiency of the pooled testing procedure. One also needs to measure the sensitivity and specificity of the pooled testing procedure, and most importantly, the probabilities of the diagnostic errors.

Let *T*^+^ and *T*^−^ denote the events that an individual is tested positive and negative, respectively, by the pooled testing procedure. Further, let *I* (*I^c^*) denote the events that an individual is infected (uninfected). Then one can easily check that the sensitivity, say, 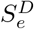, and specificity, say, 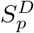, of the Dorfman’s pooled tesiting procedure, are given by

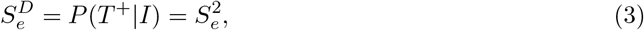

and

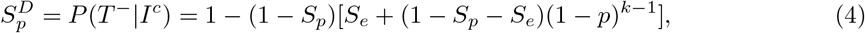

respectively. The proofs of (3)-(4) are given in the appendix.

Notice that 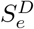 depends only on *S_e_* while 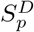 depends on all three parameters *S_e_*, *S_p_* and *p*. As stated above, 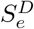 and 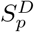 provide useful information about the performance of the pooled testing procedure. However, the posterior or inverse probabilities *P*(*I^c^*|*T*+) and *P*(*I*|*T*^−^), called the False Positive Predictive Value (FPPV) and the False Negative Predictive Value (FNPV) respectively, are often critical for assessing the performance of the test in the real life situation. These are measures of diagnostic errors. The *FPPV* and the *FNPV* represent the proportion of misclassified individuals among those who are tested positive, and among those who are tested negative, respectively. In the context of testing for sera samples of HIV virus, Litvak (^22^) proposed that the five key elements to be used to capture the overall performance of a pooled testing procedure should be: *E*(*N_j_*), 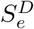, 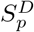, *FPPV* and *FNPV*.

Now, using Bayes’ Theorem (^20^), one can easily show that

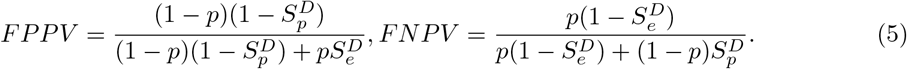

Notice that, for individual testing *FPPV* and *FNPV*, denoted henceforth *FPPV*_I_ and *FNPV*_I_ respectively, are obtained by substituting 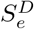 and 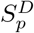 by *S_e_* and *S_p_*, respectively, in (5).

In Tables 1a and 1b, we furnish the values of *E*(*N_j_*), 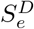, 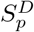, *FPPV*, *FNPV*, *FPPV*_I_ and *FNPV*_I_ for some appropriately chosen values of the sensitivity (*S_e_*), the specificity (*S_p_*) and the prevalence (*p*). The “Eff” (efficiency) column gives the percentage reduction in the expected number of tests achieved by using the pooled testing than the individual testing. Evidently, with increase in *p*, and decrease in *S_e_* and *S_p_*, the efficiency reduces. The value of 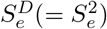 is always less than, or equal to, *S_e_*. However, for a given set of values of *p*, *S_e_* and *S_p_*, the value of 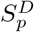 is always more than *S_p_*, and for smaller values of *S_p_* this effect is substantial. Thus, the pooled testing procedure may lead to a significant improvement of the specificity, especially when the specificity of the individual test is low. Further, it is interesting to observe that even a slight decrease in *S_p_* leads to a substantial increase in the value of *FPPV* for any given values of *p* and *S_e_*. On the other hand, a change in the value of *S_e_* has negligible effect on the value of *FPPV* for any given values of *p* and *S_p_*. Also, with increase in *p*, *FPPV* decreases for any given values of *S_e_* and *S_p_*. Most importantly, compared to the individual testing, the pooled testing leads to a substantial reduction in the value of *FPPV*, which is extremely important from the point of view of its application in practice. Still, *FPPV* can take very high values even for pooled testing for a low prevalence disease with low specificity (*S_p_*). While this is undesirable, this compares favourably to individual testing. In contrast, for the range of values considered in the tables for *p*, *S_e_* and *S_p_*, the impact of changes in the values of these parameters has little effect on *FNPV*.

**Table 1a:** Optimal group size (*k*^*^), efficiency in terms of reduction in test numbers as percentage of number of individuals tested (Eff), pooled testing sensitivity 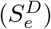 and specificity 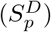 false positive predictive value (*FPPV*) and false negative predictive value (*FNPV*) corresponding to different prevalence rates (*p*), sensitivities (*S_e_*) and specificities (*S_p_*) for Dorfman’s algorithm. False positive predictive value (*FPPV_I_*) and false negative predictive value (*FWPV_I_*) for individual tests are given within parentheses to facilitate comparison.

| Test Quality | $p$ | $k^*$ | Eff (%) | $S_e^D$ | $S_p^D$ | $FPPV$ ($FPPV_I$) | $FNPV$ ($FNPV_I$) |
| --- | --- | --- | --- | --- | --- | --- | --- |
| $S_e = 1$<br>$S_p = 1$ | 0.01 | 11 | 80.44 | 1 | 1 | 0 (0) | 0 (0) |
|  | 0.02 | 8 | 72.58 | 1 | 1 | 0 (0) | 0 (0) |
|  | 0.05 | 5 | 57.38 | 1 | 1 | 0 (0) | 0 (0) |
|  | 0.10 | 4 | 40.61 | 1 | 1 | 0 (0) | 0 (0) |
| $S_e = 0.99$<br>$S_p = 0.99$ | 0.01 | 11 | 79.65 | 0.98 | 0.999 | 0.09 (0.50) | 0.0002 (0.0001) |
|  | 0.02 | 8 | 71.87 | 0.98 | 0.999 | 0.07 (0.33) | 0.0004 (0.0002) |
|  | 0.05 | 5 | 56.83 | 0.98 | 0.998 | 0.04 (0.16) | 0.0010 (0.0005) |
|  | 0.10 | 4 | 40.30 | 0.98 | 0.997 | 0.02 (0.08) | 0.0022 (0.0011) |
| $S_e = 0.99$<br>$S_p = 0.95$ | 0.01 | 11 | 76.07 | 0.98 | 0.993 | 0.41 (0.83) | 0.0002 (0.0001) |
|  | 0.02 | 8 | 68.47 | 0.98 | 0.991 | 0.30 (0.71) | 0.0004 (0.0002) |
|  | 0.05 | 5 | 53.74 | 0.98 | 0.989 | 0.18 (0.49) | 0.0011 (0.0006) |
|  | 0.10 | 4 | 37.67 | 0.98 | 0.985 | 0.12 (0.31) | 0.0022 (0.0012) |
| $S_e = 0.99$<br>$S_p = 0.90$ | 0.01 | 11 | 71.59 | 0.98 | 0.981 | 0.65 (0.91) | 0.0002 (0.0001) |
|  | 0.02 | 8 | 64.22 | 0.98 | 0.978 | 0.52 (0.83) | 0.0004 (0.0002) |
|  | 0.05 | 5 | 49.87 | 0.98 | 0.973 | 0.34 (0.66) | 0.0011 (0.0006) |
|  | 0.10 | 4 | 34.39 | 0.98 | 0.966 | 0.24 (0.48) | 0.0023 (0.0012) |
| $S_e = 0.99$<br>$S_p = 0.80$ | 0.01 | 12 | 62.69 | 0.98 | 0.943 | 0.85 (0.95) | 0.0002 (0.0001) |
|  | 0.02 | 9 | 55.75 | 0.98 | 0.936 | 0.76 (0.91) | 0.0004 (0.0003) |
|  | 0.05 | 6 | 42.41 | 0.98 | 0.924 | 0.59 (0.79) | 0.0011 (0.0007) |
|  | 0.10 | 4 | 27.83 | 0.98 | 0.917 | 0.43 (0.65) | 0.0024 (0.0014) |
| $S_e = 0.99$<br>$S_p = 0.70$ | 0.01 | 13 | 53.86 | 0.98 | 0.886 | 0.92 (0.97) | 0.0002 (0.0001) |
|  | 0.02 | 9 | 47.42 | 0.98 | 0.879 | 0.86 (0.94) | 0.0005 (0.0003) |
|  | 0.05 | 6 | 35.05 | 0.98 | 0.863 | 0.73 (0.85) | 0.0012 (0.0008) |
|  | 0.10 | 5 | 21.74 | 0.98 | 0.839 | 0.60 (0.73) | 0.0026 (0.0016) |
| $S_e = 0.95$<br>$S_p = 0.99$ | 0.01 | 11 | 80.07 | 0.90 | 0.999 | 0.10 (0.51) | 0.0010 (0.0005) |
|  | 0.02 | 8 | 72.47 | 0.90 | 0.999 | 0.07 (0.34) | 0.0020 (0.0010) |
|  | 0.05 | 5 | 57.74 | 0.90 | 0.998 | 0.04 (0.17) | 0.0051 (0.0027) |
|  | 0.10 | 4 | 41.67 | 0.90 | 0.997 | 0.03 (0.09) | 0.0107 (0.0056) |
| $S_e = 0.95$<br>$S_p = 0.95$ | 0.01 | 11 | 76.49 | 0.90 | 0.993 | 0.43 (0.84) | 0.0010 (0.0005) |
|  | 0.02 | 8 | 69.07 | 0.90 | 0.992 | 0.31 (0.72) | 0.0020 (0.0011) |
|  | 0.05 | 5 | 54.64 | 0.90 | 0.989 | 0.19 (0.50) | 0.0052 (0.0028) |
|  | 0.10 | 4 | 39.05 | 0.90 | 0.985 | 0.13 (0.32) | 0.0109 (0.0058) |
| $S_e = 0.95$<br>$S_p = 0.90$ | 0.01 | 11 | 72.01 | 0.90 | 0.982 | 0.67 (0.91) | 0.0010 (0.0006) |
|  | 0.02 | 8 | 64.81 | 0.90 | 0.979 | 0.54 (0.84) | 0.0020 (0.0011) |
|  | 0.05 | 6 | 50.82 | 0.90 | 0.971 | 0.38 (0.67) | 0.0053 (0.0029) |
|  | 0.10 | 4 | 35.77 | 0.90 | 0.967 | 0.25 (0.49) | 0.0111 (0.0061) |
| $S_e = 0.95$<br>$S_p = 0.80$ | 0.01 | 12 | 63.15 | 0.90 | 0.944 | 0.86 (0.95) | 0.0010 (0.0006) |
|  | 0.02 | 9 | 56.42 | 0.90 | 0.938 | 0.77 (0.91) | 0.0021 (0.0013) |
|  | 0.05 | 6 | 43.47 | 0.90 | 0.926 | 0.61 (0.80) | 0.0055 (0.0033) |
|  | 0.10 | 5 | 29.29 | 0.90 | 0.908 | 0.48 (0.65) | 0.0118 (0.0069) |
| $S_e = 0.95$<br>$S_p = 0.70$ | 0.01 | 13 | 54.35 | 0.90 | 0.888 | 0.92 (0.97) | 0.0011 (0.0007) |
|  | 0.02 | 10 | 48.11 | 0.90 | 0.878 | 0.87 (0.94) | 0.0023 (0.0015) |
|  | 0.05 | 6 | 36.11 | 0.90 | 0.866 | 0.74 (0.86) | 0.0059 (0.0037) |
|  | 0.10 | 5 | 23.38 | 0.90 | 0.843 | 0.61 (0.74) | 0.0127 (0.0079) |

**Table 1b:** Optimal group size (*k*\*), efficiency in terms of reduction in test numbers as percentage of number of individuals tested (Eff), pooled testing sensitivity 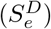 and specificity 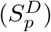 false positive predictive value (*FPPV*) and false negative predictive value (*FNPV*) corresponding to different prevalence rates (*p*), sensitivities (*S_e_*) and specificities (*S_p_*) for Dorfman’s algorithm (Continued).

| Test Quality | $p$ | $k^*$ | Eff (%) | $S_e^D$ | $S_p^D$ | $FPPV$ ($FPPV_I$) | $FNPV$ ($FNPV_I$) |
| --- | --- | --- | --- | --- | --- | --- | --- |
| $S_e = 0.90$<br>$S_p = 0.99$ | 0.01 | 11 | 80.59 | 0.81 | 0.999 | 0.10 (0.52) | 0.0019 (0.0010) |
|  | 0.02 | 8 | 73.22 | 0.81 | 0.999 | 0.07 (0.35) | 0.0039 (0.0021) |
|  | 0.05 | 5 | 58.87 | 0.81 | 0.998 | 0.04 (0.17) | 0.0099 (0.0053) |
|  | 0.10 | 4 | 43.39 | 0.81 | 0.997 | 0.03 (0.09) | 0.0207 (0.0111) |
| $S_e = 0.90$<br>$S_p = 0.95$ | 0.01 | 11 | 77.01 | 0.81 | 0.993 | 0.45 (0.85) | 0.0019 (0.0011) |
|  | 0.02 | 8 | 69.81 | 0.81 | 0.992 | 0.33 (0.73) | 0.0039 (0.0021) |
|  | 0.05 | 6 | 55.82 | 0.81 | 0.988 | 0.22 (0.51) | 0.0100 (0.0055) |
|  | 0.10 | 4 | 40.77 | 0.81 | 0.986 | 0.13 (0.33) | 0.021 (0.0116) |
| $S_e = 0.90$<br>$S_p = 0.90$ | 0.01 | 12 | 72.58 | 0.81 | 0.982 | 0.69 (0.92) | 0.0020 (0.0011) |
|  | 0.02 | 9 | 65.59 | 0.81 | 0.978 | 0.57 (0.84) | 0.0039 (0.0023) |
|  | 0.05 | 6 | 52.14 | 0.81 | 0.972 | 0.40 (0.68) | 0.0102 (0.0058) |
|  | 0.10 | 4 | 37.49 | 0.81 | 0.968 | 0.26 (0.5) | 0.0213 (0.0122) |
| $S_e = 0.90$<br>$S_p = 0.80$ | 0.01 | 13 | 63.73 | 0.81 | 0.944 | 0.87 (0.96) | 0.0020 (0.0013) |
|  | 0.02 | 9 | 57.25 | 0.81 | 0.939 | 0.79 (0.92) | 0.0041 (0.0025) |
|  | 0.05 | 6 | 44.79 | 0.81 | 0.928 | 0.63 (0.81) | 0.0107 (0.0065) |
|  | 0.10 | 5 | 31.33 | 0.81 | 0.912 | 0.49 (0.67) | 0.0226 (0.0137) |
| $S_e = 0.90$<br>$S_p = 0.70$ | 0.01 | 14 | 54.98 | 0.81 | 0.888 | 0.93 (0.97) | 0.0022 (0.0014) |
|  | 0.02 | 10 | 49.02 | 0.81 | 0.880 | 0.88 (0.94) | 0.0044 (0.0029) |
|  | 0.05 | 7 | 37.61 | 0.81 | 0.862 | 0.76 (0.86) | 0.0115 (0.0075) |
|  | 0.10 | 5 | 25.43 | 0.81 | 0.848 | 0.63 (0.75) | 0.0243 (0.0156) |
| $S_e = 0.80$<br>$S_p = 0.99$ | 0.01 | 12 | 81.69 | 0.64 | 0.999 | 0.13 (0.55) | 0.0036 (0.0020) |
|  | 0.02 | 9 | 74.75 | 0.64 | 0.999 | 0.09 (0.38) | 0.0073 (0.0041) |
|  | 0.05 | 6 | 61.41 | 0.64 | 0.998 | 0.05 (0.19) | 0.0186 (0.0105) |
|  | 0.10 | 4 | 46.83 | 0.64 | 0.998 | 0.03 (0.10) | 0.0385 (0.0220) |
| $S_e = 0.80$<br>$S_p = 0.95$ | 0.01 | 12 | 78.15 | 0.64 | 0.994 | 0.50 (0.86) | 0.0036 (0.0021) |
|  | 0.02 | 9 | 71.42 | 0.64 | 0.992 | 0.38 (0.75) | 0.0074 (0.0043) |
|  | 0.05 | 6 | 58.47 | 0.64 | 0.989 | 0.25 (0.54) | 0.0188 (0.0110) |
|  | 0.10 | 5 | 44.29 | 0.64 | 0.985 | 0.18 (0.36) | 0.0390 (0.0229) |
| $S_e = 0.80$<br>$S_p = 0.90$ | 0.01 | 13 | 73.73 | 0.64 | 0.982 | 0.74 (0.93) | 0.0037 (0.0022) |
|  | 0.02 | 9 | 67.25 | 0.64 | 0.980 | 0.61 (0.86) | 0.0074 (0.0045) |
|  | 0.05 | 6 | 54.79 | 0.64 | 0.974 | 0.43 (0.70) | 0.0191 (0.0116) |
|  | 0.10 | 5 | 41.33 | 0.64 | 0.966 | 0.32 (0.53) | 0.0398 (0.0241) |
| $S_e = 0.80$<br>$S_p = 0.80$ | 0.01 | 14 | 64.98 | 0.64 | 0.945 | 0.89 (0.96) | 0.0038 (0.0025) |
|  | 0.02 | 10 | 59.02 | 0.64 | 0.940 | 0.82 (0.92) | 0.0078 (0.0051) |
|  | 0.05 | 7 | 47.61 | 0.64 | 0.928 | 0.68 (0.83) | 0.0200 (0.0130) |
|  | 0.10 | 5 | 35.43 | 0.64 | 0.919 | 0.53 (0.69) | 0.0417 (0.0270) |
| $S_e = 0.80$<br>$S_p = 0.70$ | 0.01 | 15 | 56.34 | 0.64 | 0.890 | 0.94 (0.97) | 0.0041 (0.0029) |
|  | 0.02 | 11 | 50.95 | 0.64 | 0.883 | 0.90 (0.95) | 0.0083 (0.0058) |
|  | 0.05 | 8 | 40.67 | 0.64 | 0.865 | 0.80 (0.88) | 0.0214 (0.0148) |
|  | 0.10 | 6 | 29.91 | 0.64 | 0.849 | 0.68 (0.77) | 0.0450 (0.0308) |

Notice that in case both the sensitivity and the specificity of the test are low, say, for example, *S_e_* = 0.8 and *S_p_* = 0.7, even with 10% prevalence, 68% (77%) of the pooled (individual) testing results would be false positive, which is extremely high. A recent meta-study by a group of medical professionals from the Johns Hopkins University^30^ demonstrated that over a different varieties of RT-PCR tests, which are the most commonly used tests for SARS-CoV-2, a best case scenario is an *S_e_* ≈ 0.8. Another meta-study^31^ has looked at thirty seven different external quality assessment studies of different medical assays, and found that in some cases, the specificity was as low as 0.83.

For testing for the SARS-CoV-2 virus, the above observations suggest that at the initial phase of the pandemic when the prevalence is low, (less than 10%,) pooled testing may lead to a substantially lower rate of false positive cases than individual testing, especially if a low specificity test is used. Admittedly, pooled testing leads to a slightly higher rate of false negative cases compared to individual testing, but as observed from Tables 1a and 1b, that effect is usually negligible.

Finally, it may be noted here that the impact of the two diagnostic errors are asymmetric. In case of more false positive cases, more people are to be quarantined leading to a lot of economic, social and emotional turmoil. On the other hand, in case of more false negative results, more infected people will be released in the population causing the disease to spread faster. Considering the costs of the errors, the policy planner has to make a trade-off.

## 4 Estimation of prevalence using pooled testing data

In this section, first we present the theory of estimation of prevalence of a disease from the test data using basic probability theory^20^. Next, we study the impact of misspecification of sensitivity and specificity on the estimate of prevalence.

An accurate estimation of the prevalence of SARS-CoV-2-virus, being critical for assessing the lethality of the pandemic, is crucial for the policy makers to make strategic decisions. Recently, Ioannidis^32^, a Stanford medicine professor, lamented in a highly critical opinion piece on COVID-19: “Three months after the outbreak emerged, most countries, including the U.S., lack the ability to test a large number of people and no countries have reliable data on the prevalence of the virus in a representative random sample of the general population.” Ioannidis argued that in the absence of such data, the estimate of fatality rate is bound to be a substantial overestimate of the true fatality rate if a significant percentage of COVID-19 cases are undetected. It is now well known that a significant percentage of COVID-19 cases are asymptomatic. Raman R Gangakhedkar, chief epidemiologist, ICMR, reported “Of 100 people with infection, 80 do not have symptoms,” ^33^. A lab in Iceland^34^ has suggested that 50% of the infected are asymptomatic. In a Boston homeless shelter, out of 400 guests staying there, 146 tested positive for COVID-19, but all were reported to be asymptomatic^35^. If, for example, 80% of the cases are really asymptomatic, then the estimate of fatality rate would naturally be five times the true fatality rate. While the reported percentage of asymptomatic cases vary from place to place, all agree that it is significant. Considering the possibility of substantial underestimation of infected cases in the absence of test data, Ioannidis has contended that the virus could be less deadly than people think, and destroying the economy in the effort to fight with the virus could be a “once-in-a-century-evidence-fiasco”.

A recent research paper published by a team of Stanford medical scientists^16^ lends support to Ioannidis’ contention. By estimating the prevalence of SARS-CoV-2 virus among the residents of Santa Clara County of California using antibody test data of a “properly selected” sample of 3300 residents, it predicted that between 50 to 85 times more residents of the county were actually infected than what appeared in the official tallies^36^. The authors claimed that “their data helps prove … if undetected infections are as widespread as they think, then the death rate in the county may be less than 0.2%, about a fifth to a tenth other estimates.” The publication of this research paper immediately created waves in social media, in press, and in policy circles; if indeed, this number is not far from the truth, then the COVID-19 fatality rate is not very different from common flu. Realizing the findings’ serious policy implications, especially its lending support to the view of lifting the lock down, some scientists immediately issued a caveat about the accuracy of the estimate. They expressed concerns about the flaws in the sample selection method (through Facebook advertisements), the statistical analysis carried out, and the reliability of the antibody test^37,38^.

We now discuss the methodology used by the Stanford group^16^ for prevalence estimation based on individual serological testing data. Let us denote the probability of a positive test result for an arbitrarily chosen individual by π. Clearly, for a sample of size *n*, the estimate of π is simply the proportion of people being identified as positive out of the sample. But we know from the previous section that

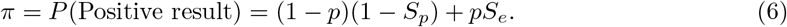

Plugging in the proportion of positive test results obtained from the sample, say 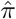, for *π* in (6), we obtain

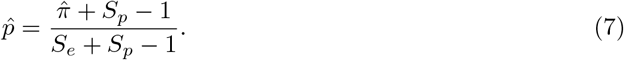

However, for estimating prevalence using Dorfman’s algorithm, we simply need to replace *S_e_* and *S_p_* in (6) by 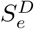 and 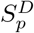 respectively (cf. (Equations 3)-(4)). Thus, we have

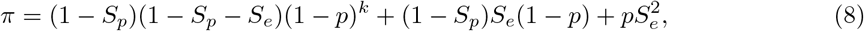

or, equivalently,

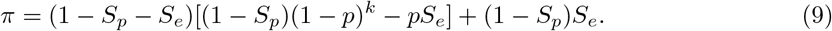

Hence, plugging in an estimate of *π* obtained from the test data to (8) and then solving it numerically for *p* we get an estimate of *p*, say 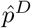. This estimate is consistent for *p* (i.e., close to *p* as the sample size *n* gets very large).

As noted in Section 3, both RT-PCR and antibody tests have high sensitivity and specificity in lab settings, but in a real world situation, these values may be substantially lower than those obtained in lab settings. Suppose 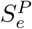 and 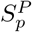 are respectively the sensitivity and the specificity values as perceived, and used by the scientist for estimating the prevalence from Equation (7) or (8), whereas 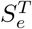 and 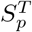 are respectively the true sensitivity and specificity values in the field. In real-life situations, it is often the case that 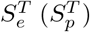 is substantially less than 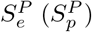, because the scientist’s perceived values are usually influenced by the values under the lab setting. Thus, given the true value of *π*, the solution to Equation (7) or (8) for *p* is equal to the true prevalence *p_T_* if *S_e_* and *S_p_* are replaced by 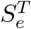 and 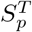. On the other hand, given the true value of *π*, replacing *S_e_* and *S_p_* by 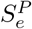 and 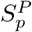 in (7) and (8), would yield a solution *p_P_* which is expected to deviate from the true prevalence *p_T_*. We study the effect of the misspecification of sensitivity and specificity values on the estimate of prevalence by evaluating the bias (*p_T_* - *p_P_*).

In Tables 3a-3b, we report the values of the bias (*p_T_* - *p_P_*) resulting from the individual testing as well as Dorfman’s pooled testing, denoted by *bias_I_* - and *bias_D_*, respectively, for different values of *p*, 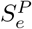, 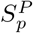, 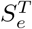 and 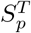. The following patterns are visible from the table:

a. With increase in the prevalence *p*, the bias is decreasing. Initially, it reduces from positive to zero and then becomes negative. We report the bias for four different values of *p*: 0.01, 0.02, 0.05 and 0.1.
b. The effect of misspecification of *S_e_* and *S_p_* are asymmetric in nature. Misspecification of *S_e_* has a little effect on the bias. However, the misspecification of *S_p_* has a significantly large effect on the bias. Also, more is the deviation from the true values more is the effect on bias.
c. The most interesting observation is, compared to the individual testing, pooled testing reduces the bias due to misspecification of *S_e_* and *S_p_* substantially. This is extremely important to know given the fact that misspecification of *S_e_* and *S_p_* are quite common.

In Table 2, we report the values of *π*, the probability of testing positive, for the individual testing as well as for the Dorfman testing procedure, for different values of *p*, *S_e_* and *S_p_*. Let *π_I_* (cf. Equation (6)) and *π_D_* (cf. Equation (8)) denote the *π* value corresponding to the individual and Dorfman testing procedure, respectively. Notice from Table 2 that, like the observation (ii) made above, the value of *π* is significantly influenced by *S_p_*, but not so by *S_e_*. Also, it is important to note that, for a given value of *S_p_* (say, 0.9) reduction in *S_e_* (say, from 1 to 0.7) results in reduction of *π*, though not a significant reduction. Thus, we may conclude that misspecification of *S_p_* (*S_e_*) has a large (negligible) effect on the value of *π*, and consequently, on the estimates of prevalence obtained by solving Equations (6) or (8), as the case may be. So, for accurate estimation of prevalence it is very important to specify the value of *S_p_* correctly, while misspecification of *S_e_* has little effect.

**Table 2:** Expected values of the proportion of sample testing positive (*π*) corresponding to different prevalence rates, sensitivities (*S_e_*) and specificities (*S_p_*) for individual testing (π*_INDIV_*) Dorfman’s algorithm (*π_DORF_*). The *π_DORF_* values are based on *k* = 4, the most conservative pooling choice from Tables 1a and 1b.

| Test Quality | p | $\pi_{INDIV}$ | $\pi_{DORF}$ | Test Quality | p | $\pi_{INDIV}$ | $\pi_{DORF}$ |
| --- | --- | --- | --- | --- | --- | --- | --- |
| $S_e = 1$<br>$S_p = 1$ | 0.01 | 0.01 | 0.01 | $S_e = 0.80$<br>$S_p = 1$ | 0.01 | 0.01 | 0.01 |
|  | 0.02 | 0.02 | 0.02 |  | 0.02 | 0.02 | 0.01 |
|  | 0.05 | 0.05 | 0.05 |  | 0.05 | 0.04 | 0.03 |
|  | 0.10 | 0.10 | 0.10 |  | 0.10 | 0.08 | 0.06 |
| $S_e = 1$<br>$S_p = 0.90$ | 0.01 | 0.11 | 0.02 | $S_e = 0.80$<br>$S_p = 0.90$ | 0.01 | 0.11 | 0.02 |
|  | 0.02 | 0.12 | 0.03 |  | 0.02 | 0.11 | 0.03 |
|  | 0.05 | 0.14 | 0.07 |  | 0.05 | 0.13 | 0.05 |
|  | 0.10 | 0.19 | 0.13 |  | 0.10 | 0.17 | 0.09 |
| $S_e = 1$<br>$S_p = 0.80$ | 0.01 | 0.21 | 0.05 | $S_e = 0.80$<br>$S_p = 0.80$ | 0.01 | 0.21 | 0.05 |
|  | 0.02 | 0.22 | 0.07 |  | 0.02 | 0.21 | 0.06 |
|  | 0.05 | 0.24 | 0.11 |  | 0.05 | 0.23 | 0.09 |
|  | 0.10 | 0.28 | 0.18 |  | 0.10 | 0.26 | 0.13 |
| $S_e = 1$<br>$S_p = 0.70$ | 0.01 | 0.31 | 0.11 | $S_e = 0.80$<br>$S_p = 0.70$ | 0.01 | 0.31 | 0.10 |
|  | 0.02 | 0.31 | 0.12 |  | 0.02 | 0.31 | 0.11 |
|  | 0.05 | 0.34 | 0.16 |  | 0.05 | 0.33 | 0.14 |
|  | 0.10 | 0.37 | 0.23 |  | 0.10 | 0.35 | 0.18 |
| $S_e = 0.90$<br>$S_p = 1$ | 0.01 | 0.01 | 0.01 | $S_e = 0.70$<br>$S_p = 1$ | 0.01 | 0.01 | 0 |
|  | 0.02 | 0.02 | 0.02 |  | 0.02 | 0.01 | 0.01 |
|  | 0.05 | 0.05 | 0.04 |  | 0.05 | 0.03 | 0.02 |
|  | 0.10 | 0.09 | 0.08 |  | 0.10 | 0.07 | 0.05 |
| $S_e = 0.90$<br>$S_p = 0.90$ | 0.01 | 0.11 | 0.02 | $S_e = 0.70$<br>$S_p = 0.90$ | 0.01 | 0.11 | 0.02 |
|  | 0.02 | 0.12 | 0.03 |  | 0.02 | 0.11 | 0.02 |
|  | 0.05 | 0.14 | 0.06 |  | 0.05 | 0.13 | 0.04 |
|  | 0.10 | 0.18 | 0.11 |  | 0.10 | 0.16 | 0.07 |
| $S_e = 0.90$<br>$S_p = 0.80$ | 0.01 | 0.21 | 0.05 | $S_e = 0.70$<br>$S_p = 0.80$ | 0.01 | 0.20 | 0.05 |
|  | 0.02 | 0.21 | 0.06 |  | 0.02 | 0.21 | 0.05 |
|  | 0.05 | 0.23 | 0.10 |  | 0.05 | 0.22 | 0.08 |
|  | 0.10 | 0.27 | 0.15 |  | 0.10 | 0.25 | 0.11 |
| $S_e = 0.90$<br>$S_p = 0.70$ | 0.01 | 0.31 | 0.10 | $S_e = 0.70$<br>$S_p = 0.70$ | 0.01 | 0.30 | 0.10 |
|  | 0.02 | 0.31 | 0.11 |  | 0.02 | 0.31 | 0.10 |
|  | 0.05 | 0.33 | 0.15 |  | 0.05 | 0.32 | 0.13 |
|  | 0.10 | 0.36 | 0.21 |  | 0.10 | 0.34 | 0.16 |

**Table 3a:** Estimation results when perceived sensitivity 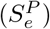 and specificity 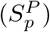 are higher than the true situations. Bias bias*_I_* corresponds to individual tests, while bias*_D_* corresponds to Dorfman tests. The Dorfman values are based on *k* = 4, the most conservative pooling choice from Tables 1a and 1b.

| Perceived test quality | p | $S_e^T = S_e^P$<br>$S_p^T$ 10% less | | $S_e^T$ 10% less<br>$S_p^T = S_p^P$ | | Both $S_e^T$ and $S_p^T$<br>10% less | | Both $S_e^T$ and $S_p^T$<br>20% less | |
| --- | --- | --- | --- | --- | --- | --- | --- | --- | --- |
| | | $bias_I$ | $bias_D$ | $bias_I$ | $bias_D$ | $bias_I$ | $bias_D$ | $bias_I$ | $bias_D$ |
| $S_e^P = 1$<br>$S_p^P = 1$ | 0.01 | 0.10 | 0.01 | 0 | 0 | 0.10 | 0.01 | 0.20 | 0.04 |
|  | 0.02 | 0.10 | 0.01 | 0 | 0 | 0.10 | 0.01 | 0.19 | 0.04 |
|  | 0.05 | 0.10 | 0.02 | -0.01 | -0.01 | 0.09 | 0.01 | 0.18 | 0.04 |
|  | 0.10 | 0.09 | 0.03 | -0.01 | -0.02 | 0.08 | 0.01 | 0.16 | 0.03 |
| $S_e^P = 0.99$<br>$S_p^P = 0.99$ | 0.01 | 0.10 | 0.01 | 0 | 0 | 0.10 | 0.01 | 0.20 | 0.04 |
|  | 0.02 | 0.10 | 0.02 | 0 | 0 | 0.10 | 0.01 | 0.20 | 0.04 |
|  | 0.05 | 0.10 | 0.02 | -0.01 | -0.01 | 0.09 | 0.01 | 0.18 | 0.04 |
|  | 0.10 | 0.09 | 0.03 | -0.01 | -0.02 | 0.08 | 0.01 | 0.16 | 0.03 |
| $S_e^P = 0.99$<br>$S_p^P = 0.95$ | 0.01 | 0.11 | 0.02 | 0 | 0 | 0.10 | 0.02 | 0.21 | 0.05 |
|  | 0.02 | 0.10 | 0.02 | 0 | 0 | 0.10 | 0.02 | 0.20 | 0.05 |
|  | 0.05 | 0.10 | 0.03 | -0.01 | -0.01 | 0.10 | 0.02 | 0.19 | 0.05 |
|  | 0.10 | 0.10 | 0.04 | -0.01 | -0.02 | 0.09 | 0.01 | 0.17 | 0.04 |
| $S_e^P = 0.99$<br>$S_p^P = 0.90$ | 0.01 | 0.11 | 0.03 | 0 | 0 | 0.11 | 0.02 | 0.22 | 0.06 |
|  | 0.02 | 0.11 | 0.03 | 0 | 0 | 0.11 | 0.02 | 0.22 | 0.06 |
|  | 0.05 | 0.11 | 0.03 | -0.01 | -0.01 | 0.10 | 0.02 | 0.20 | 0.06 |
|  | 0.10 | 0.10 | 0.04 | -0.01 | -0.02 | 0.09 | 0.02 | 0.18 | 0.05 |
| $S_e^P = 0.99$<br>$S_p^P = 0.80$ | 0.01 | 0.13 | 0.04 | 0 | 0 | 0.12 | 0.04 | 0.25 | 0.09 |
|  | 0.02 | 0.12 | 0.04 | 0 | 0 | 0.12 | 0.03 | 0.24 | 0.08 |
|  | 0.05 | 0.12 | 0.04 | -0.01 | -0.01 | 0.11 | 0.03 | 0.23 | 0.08 |
|  | 0.10 | 0.11 | 0.05 | -0.01 | -0.02 | 0.10 | 0.03 | 0.20 | 0.06 |
| $S_e^P = 0.99$<br>$S_p^P = 0.70$ | 0.01 | 0.14 | 0.05 | 0 | 0 | 0.14 | 0.05 | 0.28 | 0.11 |
|  | 0.02 | 0.14 | 0.05 | 0 | 0 | 0.14 | 0.05 | 0.28 | 0.11 |
|  | 0.05 | 0.14 | 0.05 | -0.01 | -0.01 | 0.13 | 0.04 | 0.26 | 0.10 |
|  | 0.10 | 0.13 | 0.06 | -0.01 | -0.02 | 0.12 | 0.03 | 0.23 | 0.08 |
| $S_e^P = 0.95$<br>$S_p^P = 0.99$ | 0.01 | 0.11 | 0.02 | 0 | 0 | 0.10 | 0.01 | 0.21 | 0.05 |
|  | 0.02 | 0.10 | 0.02 | 0 | 0 | 0.10 | 0.01 | 0.20 | 0.05 |
|  | 0.05 | 0.10 | 0.02 | -0.01 | -0.01 | 0.10 | 0.01 | 0.19 | 0.04 |
|  | 0.10 | 0.10 | 0.03 | -0.01 | -0.02 | 0.09 | 0.01 | 0.17 | 0.03 |
| $S_e^P = 0.95$<br>$S_p^P = 0.95$ | 0.01 | 0.11 | 0.02 | 0 | 0 | 0.11 | 0.02 | 0.22 | 0.06 |
|  | 0.02 | 0.11 | 0.02 | 0 | 0 | 0.11 | 0.02 | 0.21 | 0.06 |
|  | 0.05 | 0.11 | 0.03 | -0.01 | -0.01 | 0.10 | 0.02 | 0.20 | 0.05 |
|  | 0.10 | 0.10 | 0.04 | -0.01 | -0.02 | 0.09 | 0.01 | 0.18 | 0.04 |
| $S_e^P = 0.95$<br>$S_p^P = 0.90$ | 0.01 | 0.12 | 0.03 | 0 | 0 | 0.12 | 0.03 | 0.23 | 0.07 |
|  | 0.02 | 0.12 | 0.03 | 0 | 0 | 0.11 | 0.03 | 0.23 | 0.07 |
|  | 0.05 | 0.11 | 0.03 | -0.01 | -0.01 | 0.11 | 0.02 | 0.21 | 0.06 |
|  | 0.10 | 0.11 | 0.04 | -0.01 | -0.02 | 0.09 | 0.02 | 0.19 | 0.05 |
| $S_e^P = 0.95$<br>$S_p^P = 0.80$ | 0.01 | 0.13 | 0.04 | 0 | 0 | 0.13 | 0.04 | 0.26 | 0.09 |
|  | 0.02 | 0.13 | 0.04 | 0 | 0 | 0.13 | 0.04 | 0.26 | 0.09 |
|  | 0.05 | 0.13 | 0.04 | -0.01 | -0.01 | 0.12 | 0.03 | 0.24 | 0.08 |
|  | 0.10 | 0.12 | 0.05 | -0.01 | -0.02 | 0.11 | 0.03 | 0.21 | 0.06 |
| $S_e^P = 0.95$<br>$S_p^P = 0.70$ | 0.01 | 0.15 | 0.05 | 0 | 0 | 0.15 | 0.05 | 0.30 | 0.12 |
|  | 0.02 | 0.15 | 0.05 | 0 | 0 | 0.15 | 0.05 | 0.30 | 0.12 |
|  | 0.05 | 0.15 | 0.06 | -0.01 | -0.01 | 0.14 | 0.04 | 0.28 | 0.11 |
|  | 0.10 | 0.14 | 0.06 | -0.02 | -0.02 | 0.12 | 0.04 | 0.25 | 0.08 |

**Table 3b:** Estimation results when perceived sensitivity 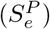 and specificity 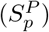 are higher than the true situations. Bias bias*_I_* corresponds to individual tests, while bias*_D_* corresponds to Dorfman tests (Continued).

| Perceived test quality | p | $S_e^T = S_e^P$<br>$S_p^T$ 10% less | | $S_e^T$ 10% less<br>$S_p^T = S_p^P$ | | Both $S_e^T$ and $S_p^T$<br>10% less | | Both $S_e^T$ and $S_p^T$<br>20% less | |
| --- | --- | --- | --- | --- | --- | --- | --- | --- | --- |
| | | $\text{bias}_I$ | $\text{bias}_D$ | $\text{bias}_I$ | $\text{bias}_D$ | $\text{bias}_I$ | $\text{bias}_D$ | $\text{bias}_I$ | $\text{bias}_D$ |
| $S_e^P = 0.90$<br>$S_p^P = 0.99$ | 0.01 | 0.11 | 0.02 | 0 | 0 | 0.11 | 0.01 | 0.22 | 0.05 |
|  | 0.02 | 0.11 | 0.02 | 0 | 0 | 0.11 | 0.01 | 0.22 | 0.05 |
|  | 0.05 | 0.11 | 0.03 | -0.01 | -0.01 | 0.10 | 0.01 | 0.20 | 0.05 |
|  | 0.10 | 0.10 | 0.04 | -0.01 | -0.02 | 0.09 | 0.01 | 0.18 | 0.04 |
| $S_e^P = 0.90$<br>$S_p^P = 0.95$ | 0.01 | 0.12 | 0.02 | 0 | 0 | 0.12 | 0.02 | 0.23 | 0.06 |
|  | 0.02 | 0.12 | 0.03 | 0 | 0 | 0.11 | 0.02 | 0.23 | 0.06 |
|  | 0.05 | 0.11 | 0.03 | -0.01 | -0.01 | 0.11 | 0.02 | 0.21 | 0.06 |
|  | 0.10 | 0.11 | 0.04 | -0.01 | -0.02 | 0.09 | 0.02 | 0.19 | 0.04 |
| $S_e^P = 0.90$<br>$S_p^P = 0.90$ | 0.01 | 0.12 | 0.03 | 0 | 0 | 0.12 | 0.03 | 0.25 | 0.08 |
|  | 0.02 | 0.12 | 0.03 | 0 | 0 | 0.12 | 0.03 | 0.24 | 0.08 |
|  | 0.05 | 0.12 | 0.04 | -0.01 | -0.01 | 0.11 | 0.03 | 0.22 | 0.07 |
|  | 0.10 | 0.11 | 0.04 | -0.01 | -0.02 | 0.10 | 0.02 | 0.20 | 0.05 |
| $S_e^P = 0.90$<br>$S_p^P = 0.80$ | 0.01 | 0.14 | 0.04 | 0 | 0 | 0.14 | 0.04 | 0.28 | 0.11 |
|  | 0.02 | 0.14 | 0.05 | 0 | 0 | 0.14 | 0.04 | 0.27 | 0.10 |
|  | 0.05 | 0.14 | 0.05 | -0.01 | -0.01 | 0.13 | 0.04 | 0.26 | 0.09 |
|  | 0.10 | 0.13 | 0.05 | -0.01 | -0.02 | 0.11 | 0.03 | 0.23 | 0.07 |
| $S_e^P = 0.90$<br>$S_p^P = 0.70$ | 0.01 | 0.16 | 0.06 | 0 | 0 | 0.16 | 0.06 | 0.33 | 0.14 |
|  | 0.02 | 0.16 | 0.06 | 0 | 0 | 0.16 | 0.06 | 0.32 | 0.13 |
|  | 0.05 | 0.16 | 0.06 | -0.01 | -0.01 | 0.15 | 0.05 | 0.30 | 0.12 |
|  | 0.10 | 0.15 | 0.07 | -0.02 | -0.02 | 0.13 | 0.04 | 0.27 | 0.09 |
| $S_e^P = 0.80$<br>$S_p^P = 0.99$ | 0.01 | 0.13 | 0.02 | 0 | 0 | 0.12 | 0.02 | 0.25 | 0.07 |
|  | 0.02 | 0.12 | 0.02 | 0 | 0 | 0.12 | 0.02 | 0.24 | 0.06 |
|  | 0.05 | 0.12 | 0.03 | -0.01 | -0.01 | 0.11 | 0.02 | 0.23 | 0.06 |
|  | 0.10 | 0.11 | 0.04 | -0.01 | -0.02 | 0.10 | 0.01 | 0.20 | 0.05 |
| $S_e^P = 0.80$<br>$S_p^P = 0.95$ | 0.01 | 0.13 | 0.03 | 0 | 0 | 0.13 | 0.03 | 0.26 | 0.08 |
|  | 0.02 | 0.13 | 0.03 | 0 | 0 | 0.13 | 0.03 | 0.26 | 0.08 |
|  | 0.05 | 0.13 | 0.04 | -0.01 | -0.01 | 0.12 | 0.02 | 0.24 | 0.07 |
|  | 0.10 | 0.12 | 0.05 | -0.01 | -0.02 | 0.11 | 0.02 | 0.21 | 0.05 |
| $S_e^P = 0.80$<br>$S_p^P = 0.90$ | 0.01 | 0.14 | 0.04 | 0 | 0 | 0.14 | 0.04 | 0.28 | 0.10 |
|  | 0.02 | 0.14 | 0.04 | 0 | 0 | 0.14 | 0.03 | 0.27 | 0.09 |
|  | 0.05 | 0.14 | 0.05 | -0.01 | -0.01 | 0.13 | 0.03 | 0.26 | 0.08 |
|  | 0.10 | 0.13 | 0.05 | -0.01 | -0.02 | 0.11 | 0.03 | 0.23 | 0.07 |
| $S_e^P = 0.80$<br>$S_p^P = 0.80$ | 0.01 | 0.16 | 0.06 | 0 | 0 | 0.16 | 0.05 | 0.33 | 0.13 |
|  | 0.02 | 0.16 | 0.06 | 0 | 0 | 0.16 | 0.05 | 0.32 | 0.13 |
|  | 0.05 | 0.16 | 0.06 | -0.01 | -0.01 | 0.15 | 0.05 | 0.30 | 0.11 |
|  | 0.10 | 0.15 | 0.07 | -0.02 | -0.02 | 0.13 | 0.04 | 0.27 | 0.09 |
| $S_e^P = 0.80$<br>$S_p^P = 0.70$ | 0.01 | 0.20 | 0.08 | 0 | 0 | 0.20 | 0.07 | 0.39 | 0.18 |
|  | 0.02 | 0.20 | 0.08 | 0 | 0 | 0.19 | 0.07 | 0.38 | 0.18 |
|  | 0.05 | 0.19 | 0.08 | -0.01 | -0.01 | 0.18 | 0.06 | 0.36 | 0.16 |
|  | 0.10 | 0.18 | 0.08 | -0.02 | -0.03 | 0.16 | 0.05 | 0.32 | 0.12 |

Finally, we explain a subtle connection between the numbers reported in Tables 3a-3b and Table 2 with a specific example. Consider the case when prevalence *p* is equal to 0.01, and the true sensitivity 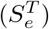 and specificity 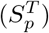 are both equal to 0.9. From Table 2, we observe that the corresponding value of *π* (cf. Equation (6)) for individual testing is equal to 0.11. Suppose now the perceived sensitivity 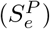 and specificity 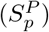 are equal to 1. To obtain the biased prevalence *p* corresponding to the perceived sensitivity and specificity, we plug in 0.11 for *π* on the left hand side of Equation (6), and *S_e_* = *S_p_* = 1 on the right hand side. The solution can be obtained from Table 2 by observing the value of *p* corresponding to *S_e_* = *S_p_* = 1 and *π_I_* = 0.11, which is clearly 0.11. Thus the resulting bias due to misspecification is 0.11 − 0.01 = 0.10, which is reported in Table 3a. By using a similar argument we can find the bias for pooled testing which is 0.01 using Table 2.

## 5 Concluding Remarks

In this article, we have discussed the statistical theory behind Dorfman’s pooled testing technique used for screening. We have explained the method of estimation of prevalence from individual testing and pooled testing data. Most importantly, we have provided theoretical insights into the practical issues that are being discussed in the scientific community, arising out of the fact that the tests for the SARS-CoV-2 virus are imperfect. The theoretical results show that pooled testing is not only preferable for reducing time and cost of screening, it also helps in significant reduction of misclassification among those who are tested positive. Our results also show that for prevalence estimation, pooled testing is always preferable to individual testing, especially when the specificity of the test is low. It helps in reducing the bias of the prevalence estimate significantly. Finally, it is worth mentioning that our observations are valid for low values of prevalence, possibly less than or equal to 10%.

In this connection, as a final remark, we may conclude that for estimating the prevalence of SARS-CoV-2 of Santa Clara County, California^16^, the Stanford scientists collected data by conducting individual antibody tests on the 3300 subjects. Instead, had they used pooled testing technique, it would have been possible to collect the test data from a much larger sample for a similar cost and time. Also, as mentioned in the paper, given that the prevalence is low (estimated as 2.8%), adopting pooled testing technique would have made all the more sense in the context of their study.

## Data Availability

No data used.

## Appendix

### A Computation of 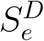 and 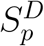

In the context of the Dorfman test, 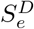, the probability that a positive person can come positive is the the probability that both the pooled test as well as the individual test comes correct, which is 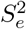.

To compute the misclassification probability that a person is non-diseased and is found to be diseased, we first note that a non-diseased person, say Person *j*, can either be a part of a pool constituted entirely of non-diseased individuals (Case A) or a pool containing some diseased individuals (Case B). The probability of Case A is (1 − *p*)*^k^*, and the probability of Case B is (1 − *p*) − (1 − *p*)*^k^*.

Now, for Person *j* to be found to be diseased in Case A, the pooled test, and subsequently the individual’s test, will both erroneously give positive result, the probability of which is (1 − *S_p_*)^2^ for the original Dorfman algorithm. On the other hand, in Case B, the pooled test will be positive, but this time correctly, while the individual’s test will still be erroneously positive, the probability of which is *S_e_*(1 − *S_p_*). Putting all together, the probability that a non-diseased person is found to be diseased is [(1 − *S_p_*)^2^(1 − *p*)*^k^* + *S_e_*(1 − *S_p_*)((1 − *p*) − (1 − *p*)*^k^*))]/(1 − *p*) = (1 − *S_p_*)*S_e_* + (1 − *Sp*)(1 − *S_p_* − *S_e_*)(1 − *p*)*^k^*^−1^. Hence, 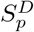 is 1 − (1 − *S_p_*)*S_e_* + (1 − *S_p_*)(1 − *S_p_* − *S_e_*)(1 − *p*)*^k^*^−1^.

